# Early impacts of the COVID-19 pandemic on mental health care and on people with mental health conditions: framework synthesis of international experiences and responses

**DOI:** 10.1101/2020.06.15.20129411

**Authors:** Luke Sheridan Rains, Sonia Johnson, Phoebe Barnett, Thomas Steare, Justin J Needle, Sarah Carr, Billie Lever Taylor, Francesca Bentivegna, Julian Edbrooke-Childs, Hannah Rachel Scott, Jessica Rees, Prisha Shah, Jo Lomani, Beverley Chipp, Nick Barber, Zainab Dedat, Sian Oram, Nicola Morant, Alan Simpson, COVID-19 Mental Health Policy Research Unit Group

**Author notes:** LSR and SJ are joint first authors. JL and BC are authors of the Lived Experience Commentary which follows the Conclusion, with contributions from SC, PS and NB.

## Abstract

**Purpose:** The COVID-19 pandemic has many potential impacts on people with mental health conditions and on mental health care, including direct consequences of infection, effects of infection control measures and subsequent societal changes. We aimed to map early impacts of the pandemic on people with pre-existing mental health conditions and services they use, and to identify individual and service-level strategies adopted to manage these.

**Methods:** We searched for relevant material in the public domain published before 30 April 2020, including papers in scientific and professional journals, published first person accounts, media articles, and publications by governments, charities and professional associations. Search languages were English, French, German, Italian, Spanish, and Mandarin Chinese. Relevant content was retrieved and summarised via a rapid qualitative framework synthesis approach.

**Results:** We found 872 eligible sources from 29 countries. Most documented observations and experiences rather than reporting research data. We found many reports of deteriorations in symptoms, and of impacts of loneliness and social isolation and of lack of access to services and resources, but sometimes also of resilience, effective self-management and peer support. Immediate service challenges related to controlling infection, especially in inpatient and residential settings, and establishing remote working, especially in the community. We summarise reports of swiftly implemented adaptations and innovations, but also of pressing ethical challenges and concerns for the future.

**Conclusion:** Our analysis captures the range of stakeholder perspectives and experiences publicly reported in the early stages of the COVID-19 pandemic in several countries. We identify potential foci for service planning and research.

## Background

The COVID-19 pandemic, declared by WHO on 11th March 2020, has health and social consequences with few peacetime precedents. The immediate focus of research and policy has understandably been on direct control of the outbreak and its repercussions for frontline staff.^1,2^ Scientific attention has also focused on the psychological wellbeing of the general population,^3,4^ and potential negative effects of quarantine and of infection.^5,6^

Less attention has however been paid to consequences for people with pre-existing mental health problems, and for the mental health services they use. It is hypothesised that they may be disproportionately negatively affected because they area already more likely to be experiencing social isolation and exclusion, stigma, and financial, employment and housing difficulties.^7,8^

Potential short-term impacts on people with pre-existing mental health conditions include:

- Effects of being infected with COVID-19, including any psychiatric sequelae, potentially increased risk of being infected or of severe COVID-19 among some groups of people with mental health conditions, and concerns regarding equitable provision of physical healthcare.
- Effects on people with mental health problems resulting from infection control measures, including potential impacts of social isolation, and lack of access to usual supports, activities and community resources.^8^
- Challenges associated with infection control in group settings, especially in hospitals and residential settings.
- The effects of reduced or re-configured mental health care delivery.

Various adaptations and innovations to enable mental health services to respond to new requirements have been discussed, including infection control strategies on mental health service premises, and remote working.^9,10^ While a number of position papers have been published, there has been relatively little systematic documentation of the impacts of the pandemic on people already living with mental health problems and on mental health care, and of strategies to mitigate these. These have been identified as urgent priority research areas.^7,8^ We aim to begin addressing this by searching for and summarising relevant material in the public domain early in the pandemic, including accounts published by people with relevant lived experience, practitioners, mental health organisations and policy makers, and also by journalists who have investigated experiences and perspectives of service users, carers and service providers.

## Aims and methods

Our aim was to conduct a document analysis to create an initial mapping and synthesis of reports, from a number of perspectives, on the early impacts of and responses to the COVID-19 pandemic on mental health care and people with mental health conditions. We drew on published sources of all types and included several languages used in countries in which the impact of the pandemic has been severe. We conducted a framework synthesis to summarise themes from sources reporting narratives and experiences of the impact of the pandemic and describing responses to it at both individual and service levels. We had planned, if warranted, also to conduct a narrative synthesis of any scientific data retrieved in our search: there was little as yet (Table 2a): this paper therefore primarily reports on our analysis of a wide range of documents in the public domain through a rapid qualitative framework synthesis method.

Specifically, we sought to analyse reports regarding:

- Direct impacts of COVID-19, subsequent public health measures and sudden social changes on people with pre-existing mental health conditions and their families.
- Self-help and informal help strategies utilised by service users and carers
- Challenges faced by mental health services during the pandemic.
- Innovations and adaptations to mitigate impacts of COVID-19 on mental health services, and reports regarding their effectiveness.

Rapid syntheses are recommended by the World Health Organization as an appropriate and timely method in rapidly developing situations,^11^ quickly providing actionable evidence that can help inform health system responses.

Our protocol was prospectively registered on PROSPERO (CRD42020182182).

## Search Strategy

We took a multi-faceted approach, in order to scope a broad, rapidly updating literature base and identify reports, articles and media from a wide range of perspectives and countries. The following database and ‘grey’ literature searches were conducted:

1. Search of four bibliographic databases (PsycINFO, PubMed, Social Science Citation Index, CINAHL) for any published scientific literature (01/01/20-17/04/20).
2. A search of relevant journal, professional body, governmental and mental health organisation websites (01/01/20-30/04/20).
3. A web search of Google Advanced, DevonAGENT Express and Blogsearchengine.org (01/01/20-30/04/20) for relevant articles and news reports published on organisational websites.
4. A Google News search for additional news articles (01/04/20-30/04/20).
5. Expert recommendations were sought throughout April, primarily from the Mental Health Policy Research Unit researcher network, and we also retrieved eligible articles from Twitter links.
6. Google searches using the same search terms as the English search (3, 4), translated by native German, French, Italian, Spanish and Mandarin speakers.
7. Searches of relevant mental health organisation websites in the study languages.

The detailed terms used for database and web searches appear at the end of the Supplementary Report (supplementary report, section 2).

## Selection Criteria

We included items meeting the following criteria:

- ***Population***: Mental health services, people using any mental health services or mental health service staff.
- ***Phenomena***: COVID-19.
- ***Focus***: Relevant to at least one of the topics above, focusing on people already living with mental health conditions at the onset of the pandemic, or on mental health care.
- ***Source type***: Published paper, article, blog post commentary, online media (including videos and podcasts), relevant to the research questions. Social media were excluded, as were blogs and articles not published via a public media channel or on the website of a public body or charity.
- ***Date***: January 2020-April 2020.
- ***Language***: Publications in English, French, Spanish, German, Italian or Mandarin Chinese. We selected these languages as we were able to involve native speakers and anticipated a substantial relevant literature.

We excluded items focusing mainly on learning disability, autism or dementia, unless combined with comorbid mental health problems, and those mainly discussing staff wellbeing. Queries were raised with the wider research team and discussed until consensus was reached. A senior reviewer (SJ) checked a subset of articles to validate inclusion decisions.

## Data extraction and analysis

For each included item, title, author, website address, access date, source type (e.g. journal article, news report, video, guidance, etc.), country, setting, service user group, and author background(s) were extracted.

Framework-based synthesis was used to enable a systematic and structured approach to rapidly summarising and analysing a large dataset.^12,13^ Analysis comprised the following steps:

### Familiarisation and development of an analytical framework

Three researchers (SJ, LSR, TS) familiarised themselves with relevant materials of various types, and then developed an initial analytic framework, comprising questions related to our study topics. We developed a semi-structured data collection form using Qualtrics software (Qualtrics, Provo, UT) to capture data from each article (supplementary report, section 3). Eight researchers piloted the form with five articles, and the framework was adjusted and finalised based on their coding and feedback. “Other” and “research reflections” categories were included to allow capture of material not covered by the initial framework.

### Indexing and charting

Included materials from the search were indexed and charted using the online form. To conduct this work rapidly in all included languages, a large number of researchers (n=62) were involved, most postgraduates, research staff or lived experience researchers linked to University College London or King’s College London. Examples of well-coded articles were provided, and the first articles coded were checked (by LSR, BLT, TS) to ensure consistency, with further training for individual researchers provided as necessary. These data were then imported into Microsoft Excel to create the framework matrix for mapping and interpretation.

### Mapping and interpretation

Twelve researchers experienced in qualitative analysis (SJ, TS, LSR, PB, JN, BLT, FB, JEC, HS, JR, PS, SC) mapped, interpreted, and summarised the data. Initially, a thematic framework was developed through discussion among them and with senior study researchers. Each researcher was assigned a portion of the data (normally one or more themes) and asked to summarise all data in the framework matrix relevant to that theme into narrative and tabular summaries. They returned to original sources if summaries were unclear or too limited. These were then discussed, further synthesized and combined to produce the results below and in the Supplementary Report (Section 1).

## Findings

We found 872 relevant sources, including 22 articles from the published literature databases, 84 from web searches focused on relevant organisations, and 266 from search engines: sources are listed in the linked Mendeley repository.^14^ 350 non-English language articles were identified through translated searches. 150 further articles were identified through expert recommendation and tweets. Included articles were English (N=503), Chinese (N=24), French (N=108), German (N=94), Italian (N=99) and Spanish (N=44). Table 1 summarises included article characteristics.

**Table 1.** Characteristics of included sources.

|  | **N (%)** |
| --- | --- |
| **Country** | Total = 872 |
| Europe | 583 (66.8) |
| UK | 250 (28.7) |
| France | 88 (10.1) |
| Germany | 79 (9.1) |
| Italy | 102 (11.7) |
| Spain | 34 (3.9) |
| Switzerland | 16 (1.8) |
| Other European countries (4) | 14 (1.6) |
| North America | 153 (17.5) |
| USA | 136 (15.6) |
| Canada | 11 (1.3) |
| Other North American countries (4) | 6 (0.7) |
| South America (3) | 5 (0.6) |
| Africa (2) | 3 (0.3) |
| Asia |  |
| China | 40 (4.6) |
| Other Asian countries (3) | 4 (0.5) |
| Australasia (2) | 10 (1.1) |
| International | 70 (8) |
| **Language** | Total = 872 |
| English | 503 (57.7) |
| Chinese | 24 (2.8) |
| French | 108 (12.4) |
| German | 94 (10.8) |
| Italian | 99 (11.4) |
| Spanish | 44 (5) |
| **Source type** | Total = 872 |
| Journal articles | 74 (8.5) |
| Media article | 442 (50.7) |
| Organisation website | 174 (20) |
| Policy document/Official guidance | 30 (3.4) |
| Public/Open letter | 7 (0.8) |
| Published blog | 105 (12) |
| Report | 7 (0.8) |
| Video/Webinar/Podcast | 33 (3.8) |
| **Author type** |  |
| Journalist | 372 |
| Clinicians and practitioners | 186 |
| People with relevant lived experience | 102 |
| Policy, professional and charity sector | 134 |
| bodies | 102 |
| Scientist | 9 |
| Other | 58 |
| Unknown |  |

Detailed summaries for each theme are in our Supplementary Report (section 1): here we provide an overview. We focus firstly on reports regarding impacts of the pandemic on people living with mental health problems and individual strategies for coping, then on impacts on the mental health care system and adaptations and innovations put in place. Similar themes appeared to emerge across countries and types of source, so all are reported together. Any peer-reviewed papers reporting data are identified below; position papers, editorials and other work describing perspectives and experiences of scientists rather than data are synthesised along with other sources.

## Impacts on mental health of people with pre-existing conditions (Supplementary Report Tables 2 and 2a)

Most sources on this topic gave narratives and personal observations regarding deteriorating mental health (supplementary report, table 2), but a handful of surveys had been conducted among people with mental health conditions (supplementary report, table 2a). A survey of young people with mental health needs, and two surveys of adults with mental health problems, all carried out for UK mental health charities, found that around four out of five respondents described experiencing increased mental health difficulties following the onset of the pandemic.^15-17^ The survey of young people reported high levels of anxiety and impulses to self-harm in the week in which schools closed in England. A report from a survey focused on financial impacts elicited self-reports of poorer wellbeing in adults with mental illness being linked to current financial and employment concerns.^18^ A further survey carried out by an academic organisation and a charity again elicited many self-reports of worsened mental health very early in the pandemic.^19^ A USA research study in pre-print showed self-reports of worse mental health in the majority of adults with mental illnesses, with only approximately one in ten feeling that they were coping well with the situation.^20^ In a small published Chinese study, Hao and colleagues found that service users were experiencing more severe mental health symptoms than the general population at the peak of the crisis in China.^21^ We found no longitudinal surveys, and only the small Chinese study included a control group.

Many sources reported observations from clinicians or self-reports of negative impacts on pre-existing mental health conditions. Mechanisms suggested for this included increased anxiety and fear of illness and death directly related to COVID-19; impacts of “lockdowns” and social distancing policies, especially of isolation; interactions between symptoms of mental health problems and current public events and concerns; impacts of loss of support from health and other services; and effects of increased social adversities, such as domestic abuse, family conflict or loss of employment.

Some accounts described impacts on specific mental health conditions, while others simply described an overall negative impact. Some conditions have been the focus of numerous and detailed narratives. Among people with depression and anxiety, sudden loss of the routines and activities that help people keep well, loneliness and isolation, and increases in health anxiety related to COVID-19 are recurrently identified as exacerbating factors. Many articles on obsessive compulsive disorder (OCD) described struggles with requirements for hygiene that contradict usual strategies for managing OCD and intensification of obsessional thoughts about contamination of infection. Regarding people with eating disorders, we found many reports that loss of eating and social routines, disrupted access to food and the increased societal prominence of food seem to exacerbate some people’s symptoms. One survey of 32 service users with eating disorders found that 38% reported worsening of symptoms during the first two weeks of lockdown in Spain.^22^ Negative impacts on mental health conditions were described alongside resilience in adversity and even some positive experiences of the pandemic period (see below).

Several scientific and media articles predicted a rise in suicide. However, an international collaboration of suicide experts argued this should not be accepted as an inevitability, but mitigated through urgent development of suicide prevention strategies.^23^ A few sources also described exacerbation of mental health problems as people replace usual coping strategies with more problematic ones, such as alcohol and substance use or gambling.

## Experiences of people with mental health problems

The pandemic has resulted in extensive and sudden social changes and new risks, some with particular relevance to people living with mental health problems. We mapped the following themes:

### Loneliness and isolation (Supplementary Report Table 3)

Many sources described loneliness, social isolation and loss of usual activities, and the negative impacts of these on mental health. Many people with mental health problems rely on the stability of routines and social contacts to manage their mental health condition, feel connected, and detect signs of deterioration. Loneliness was described as arising both from general restrictions on activities and contacts, and from sudden closure of services, including therapeutic sessions and groups, which had been sources of highly valued contacts. Patients in inpatient settings have been particularly affected by suspension of visits and leave, sometimes leading to extreme isolation and loneliness, especially when compounded by requirements to stay in hospital rooms and cancellation of ward activities.

### Lack of access to essential services and resources (Supplementary Report Table 4)

Negative impacts from closure or restriction of a range of services were frequently discussed (see also below for discussion regarding service-level changes). Some individuals reported abrupt termination or interruption of their treatment, or the replacement of face-to-face appointments by brief check-in phone calls. Others reported being unable to access care for new difficulties, or the postponement of periods of psychological therapy that were about to begin. Some sources described feeling abandoned, with a lack of access to information about how to seek urgent help if needed or about when care might resume. Remote care was not always seen as sufficient, due to lack of access to or ability to use technology, lack of privacy to engage in remote appointments, and more superficial therapeutic contacts. Interruption to medication access and adherence was also reported by several sources, including disruptions to supply or to in-person contacts required to prescribe, monitor side effects and toxicity, and administer medication. Some sources reported deterioration in mental health in the context of cessation of medication or lack of monitoring or care.

A common theme was that “we are not all in this together”, with COVID-19 risks magnifying existing inequalities and creating new ones.^24^ Thus COVID-19 and accompanying restrictions were seen by some sources as disproportionately affecting those already experiencing health and social inequalities, through economic impacts, the greater hardships of social restrictions in poor living circumstances, and the withdrawal or restriction of services disproportionately relied on by more deprived populations.

### Family and social adversities, safeguarding (Supplementary Report Table 5)

Withdrawal or reduction of services has been described as resulting in substantially more pressure for families and carers to support service users and manage distress and behavioural difficulties. Some families with caring responsibilities have reported feeling abandoned by services, especially in the context of the stresses and greater isolation associated with the “lockdown”. Meanwhile, some service user accounts expressed worry about ‘burdening’ relatives by relying on them during the lockdown, or about risks of infecting relatives with COVID-19, particularly those at greater risk of severe illness. There were also some positive descriptions of enhanced relationships with family and friends during this period, especially by keeping in touch more online or by phone, and some had moved in with family and become closer as a result.

A widely expressed concern regarding families shut in together related to the risk of increased conflict, aggression, and violence between household members and especially towards children: many sources expressed concern about this, while a smaller number described relevant incidents. Concerns related to people with mental health problems both as victims and as perpetrators. The advice to “stay home” is challenging when home is not a safe space. Both current household circumstances and reduced access to police, social services, schools and courts are identified as risk factors for continuing conflict and abuse. Seeking help may be difficult if abusers are in constant proximity. Sources argued that systems of care and outreach need to be provided for at-risk populations, potentially including communication of these via social media.

### COVID infection risks (Supplementary Report Table 6)

We did not find sources on the extent of COVID-19 infection among people with mental health problems, or whether rates of infection, or of severe consequences of infection, differ from the general population, nor were there many individual narratives regarding the experience of COVID-19 infection among people with mental health problems. There were some accounts of outbreaks of infection in hospital and residential settings and of service problems that might contribute to these, for example in the USA, China and Italy (see below regarding inpatient service challenges).

Many authors noted that co-morbidity between mental and physical health problems, and lifestyle factors (drug and alcohol use, obesity or, in the case of eating disorders, malnutrition), may result in potentially greater risk of infection and severe infection. Particular concerns were raised regarding people living in poor housing and confined, crowded, or chaotic environments, such as prisons, inpatient or residential settings, or the homeless mentally ill, as hygiene, infection control, and physical distancing practices are likely to be especially challenging. Some reports relate to people individuals with mental health problems experiencing “dual stigma” in terms of additional barriers to accessing physical healthcare: concerns related to quality of treatment for COVID-19 infection in psychiatric hospitals are discussed below.

### Positive experiences of life during the pandemic (Supplementary Report Table 7)

While negative reports exceeded them, some positive aspects of life during the pandemic were described in first person accounts, and via clinicians. Some people drew comfort from feeling that everyone was ‘in the same boat’: that people were experiencing a ‘shared trauma’ or that the rest of society was now experiencing similar challenges to the ones they faced day-to-day, such as social isolation or anxiety, and so have greater empathy. Feelings of decreased marginalisation, greater acceptance by wider society, and increased levels of community and solidarity were reported. For others, the focus on the pandemic distracted them from their pre-existing conditions, with some reporting fewer symptoms.

Secondly, some described being able to mobilise existing reserves of resilience and coping skills during the pandemic, sometimes resulting in an increased confidence. Finally, there were many reports of people taking advantage of innovations in remote and digital support, and the increasingly widespread use of video calls for communication, support and social contact. These were particularly valued by people for whom difficulties such as physical mobility, social anxiety or paranoia impede face-to-face contacts.

## Strategies people with mental health problems use to cope with the pandemic

### Individual self-management strategies (Supplementary Table 8a)

Many publications describe strategies that people with pre-existing mental health conditions have used to manage their mental health and social stresses during the pandemic. A pressing need for many has been to try to replace the activities, routines and contacts that usually support self-management. Reported self-management strategies in the pandemic have included engaging in purposeful, creative or relaxing activities, such as cooking or painting, or keeping journals to record worries or positive experiences. Use of therapeutic and self-help techniques, such as mindfulness, exposure therapy or meditation, was widely reported, though some found these of limited usefulness given current challenges. Others have sometimes found helpful self-management tools and resources, including helplines, online therapy services, websites, podcasts and apps.

The importance of maintaining a positive attitude, of self-acceptance and of not putting oneself under pressure was widely expressed. Looking after one’s physical health, such as taking regular exercise and healthy eating, maintaining a daily routine, and keeping in contact with trusted friends and family members, was emphasised in many sources. A number of people, particularly those with anxiety, reported attempting to avoid or substantially reduce their consumption of potentially stressful or triggering media coverage of the pandemic, relying instead on official or other trusted sources.

### Peer and community support (Supplementary Table 8b)

Several sources described types and impact of practical and emotional support among peers. This included mutual support and practical help, such as collecting medication. Sharing experiences and stories of mental health management, coping strategies and positive adaptations featured. Digital and online approaches to delivering support had been proactively and creatively deployed in some peer networks to facilitate one-to-one, group and community connections and activities (including recreation and socialising).

Communicating and connecting were considered vital for reducing social isolation in lockdown, managing mental health, and maintaining relationships with friends, family and peer support networks. The importance of connecting with others in inpatient settings during the pandemic was also mentioned. Mutual aid among peers appeared to have positive wellbeing benefits for those offering support.

## Service impacts

### Changes in service activity (Supplementary Table 9)

Reports based on official data were not generally available at this early stage, but several sources included reports from service managers and clinicians regarding service activity. Most reported reduced referrals and presentations to community mental health services, emergency departments and psychiatric wards in the early phases of the pandemic, though one Italian source described a subsequent rise. Potential explanations included service users’ fears of infection, beliefs that help would not be available, or wishes not to burden services. Meanwhile, large increases were reported in several countries in use of relevant helplines and, in the USA, a rise in prescriptions for mental health medication.

## Service challenges and adaptations

### Challenges in inpatient and residential settings (Supplementary Table 10)

In inpatient settings and supported housing where people live together, immediate concerns were with preventing the spread of infection while attempting to maintain a therapeutic environment. Regarding immediate infection control, clinicians’ reports from several countries described a lack of protective equipment, an inability or unwillingness of some patients to adhere to protocols, and difficulties with distancing due to ward and office layouts. Lack of realistic guidance specific to mental health settings was recurrently reported. Lack of expertise or facilities to treat people with COVID-19 effectively was identified as a challenge in providing equitable care, especially where pressure was reported to treat people with mental health problems and COVID-19 as far as possible within psychiatric hospitals. A tension was frequently reported between providing good quality mental health care and infection control, with many inpatients confined to their rooms much of the time with limited face-to-face contacts and little access to advocacy, group-based therapeutic activity or trips into the community.

### Service adaptations and innovations in inpatient and residential settings (Supplementary Table 11)

The most frequently reported inpatient adaptation to meet these challenges was the creation of COVID-19 specific units for psychiatric patients with confirmed or suspected illness, often with support from physical health care professionals and protocols in place for transfer to intensive care if needed. Other infection control measures included quarantine following admission, early discharge and initiatives to reduce admissions, staggered mealtimes and reduced use of communal spaces. An innovation described by several sources was enhanced use of technology to enable remote contact with healthcare professionals for therapy during hospital admissions, and with families to maintain social contact. In some settings, depending on current national restrictions, group therapy sessions and external visits were maintained with use of Personal Protective Equipment (PPE) and physical distancing protocols. Although supported housing settings face some similar challenges to inpatient units, we found few reports about these.

## Challenges in community settings

### Challenges in community settings (Supplementary Report Table 12)

The predominant challenges reported in community settings were the need to reduce face-to-face contact and to cope with reduced capacity due to staff absence, diversion of resources to COVID-19 wards, and reduced community resources in general. Settings where service users mingle (e.g. day services) tended to have closed, and in some regions, for example of Spain and Italy, all but urgent response appeared to have closed at the onset of the pandemic, diverting resources to physical healthcare. However, a more usual response around the world appears to have been maintaining community service provision, but with much more restricted face-to-face contact. For the face-to-face working that has continued, poor access to PPE and lack of clear infection control procedures featured in reports from community mental health settings in several countries.

### Service adaptation and innovations in community settings (Supplementary Table 13)

Tele-health tools appear to have been rapidly implemented in community mental health services across the globe, allowing care to continue at least to some extent. Video calls are used both for staff meetings and patient contacts, with some innovative use for group and activity programmes. The use of digital tools such as apps and websites for therapy appears to have also increased, but was less discussed. This shift to telemedicine appears to be welcomed for use in some contexts by many clinicians and service users, who expect this to outlast the pandemic. However, important impediments and limitations were that some service users lacked technological access and expertise, or privacy for calls; poor technology resources in services; and potential negative impacts on rapport and therapeutic relationships. The voices of the digitally excluded are particularly likely to remain unheard.

## Ethical challenges (Supplementary Report Table 14)

Several challenges were identified in maintaining professional values and human rights during the pandemic. These especially - although not exclusively – centred on inpatient psychiatric settings. Some sources, especially from France, argued that access to physical health care (for COVID-19) is inequitable for mental health service users, and that they may receive poorer quality health care, due to stigma and to a policy of treating them as far as possible in psychiatric units rather than general hospitals. There were also concerns that mental health care may become less ethical during the pandemic, with clinicians and service users in various countries reporting beliefs that medication doses and the use of sedation have increased, or that coercive and restrictive practices which impact rights and freedoms may be rising, especially in wards with compromised therapeutic environments and access to advocates. Though they have not as yet been put into practice, the provisions in the emergency Coronavirus Act 2020 in England and Wales were reported to have caused great concern by potentially allowing involuntary admission decisions to involve fewer healthcare professionals, extending time limits on detention and facilitating the use of treatment without consent. Reduced access to legal representation and advocacy was also reported.

## Expectations and concerns for future (Supplementary Report Table 15)

The final theme concerned fears and expectations about the future. Internationally, a delayed wave of increased need for services was widely anticipated, potentially combined with reduced resources to meet this, especially where services are already underfunded. The potential long duration and fluctuating nature of the pandemic was also a concern: coping strategies may not be sustained at individual or service levels.

## Discussion

### Main findings and implications

We summarise here the first reports regarding the impact of the COVID-19 pandemic from a wide variety of sources, mapping the impacts, concerns, experiences and responses at an early stage from a variety of perspectives and locations, focusing on recurrent themes.

Reports suggest that individuals with mental health problems have much to cope with: pandemic fears and circumstances interact with some symptoms; routines, contacts and activities that people have developed to manage their mental health have been shattered; and loneliness and social isolation are more prevalent. The risk that social adversities and existing inequalities may get worse is very concerning. While the current situation is new, these reports are congruent with findings of persisting negative psychological and socio-economic impacts arising from previous epidemics.^5,25,26^ However, the narratives we examined also caution against making assumptions about impacts, as responses to the pandemic clearly vary. Many people with mental health problems are unfortunately used to isolation and adversity: this may result in resilience and abilities to manage challenges actively and to draw on peer and community support. Initiatives that support them in this are potentially valuable.

Regarding service impacts, the immediate wave of increased activity predicted by some seems not to have occurred, or to have shifted to services such as helplines. However, a later surge of activity is widely expected. Currently, some of the most pressing concerns relate to inpatient and residential care settings. In these environments there are both specific and immediate challenges regarding infection control, with severe potential consequences for failure, and a pressing need to combine infection control with maintaining a therapeutic environment, safeguarding patient rights, and avoiding isolation in hospital. Rapid research to investigate and compare strategies to address these challenges would be valuable.

In the community, reports of telehealth having been swiftly adopted are striking given that implementation of innovations in health services is often observed to be slow:^27^ both clinician and service user responses suggest it may well endure after the pandemic. Adoption of telehealth has previously been slow in many countries, despite evidence that it can be an effective, cost-effective and acceptable approach to reducing treatment gaps and improving access to mental health care for service users, especially where access is otherwise limited.^28-30^ We suggest that an urgent task now is to further co-produce, test and implement promising telepsychiatry initiatives so that they are as effective and acceptable as possible, drawing on already available guidance and evidence.^31^ Barriers need to be addressed, the most appropriate technologies identified, and both staff and service users supported in their use. Meanwhile, the limitations of these technologies and the need to be selective in their use also need to be recognised, especially where continuing use following the pandemic is contemplated. A range of legal, regulatory, organisational and cultural challenges will also need to be addressed.^32,3^

## Limitations

Our search was wide-ranging, capturing many perspectives, but not likely to be comprehensive. We have compressed a large amount of material into a small space in order to ensure that it is useful (our Supplementary Report provides much more detail). Although we encompass multiple countries and languages, our scope is not global, and most notably includes few reports from low- or middle-income countries. People with experience of using mental health services and mental health clinicians were involved in many ways with this research, but day-to-day management was mainly by academic researchers not currently using or working in services.

We adopted a rapid qualitative process for coding and summarising the data,^34^ not including substantial double coding: experienced researchers checked each coder’s first summaries, and during the summarising process, we returned to sources where there was inconsistency or lack of clarity, but it is likely that ideas and themes were missed. Our process was primarily deductive and based on a positivist paradigm, although discussions amongst team members with qualitative analysis experience, and use of narrative summaries, helped to retain the inductive spirit of qualitative analysis within a large and rapid analytic process. As yet, relevant scientific data are few. We have grouped together narratives and observations from all other types of sources on the basis that when scientists are reporting views, experiences and predictions rather than research findings, these are not necessarily more informative than the experiences of people trying to manage their own mental health problems or of clinicians trying to support them and to maintain services. Journalists do not generally follow the same principles of objectivity as scientists, but in a rapidly evolving situation their investigations have the advantages of being quickly carried out and of often reporting on direct contacts with service users and/or clinicians. They may, however, tend to focus on more extreme situations, just as the people with lived experiences or clinicians who write about their experiences are unlikely to be representative. Their swiftly written reports do however, provide a rich and varied corpus of material through which we can understand the range of early experiences, responses, knowledge and practice among people with pre-existing conditions and in the services that they use.

## Conclusion

With this work we have created an early map of impacts and responses from the COVID-19 pandemic that identifies areas requiring service and policy response, and many potential areas for future investigation. We note, however, that the current crisis is evolving rapidly, and suggest that while some concerns are likely to be consistent, it will be essential to continue to review needs, challenges and the success of responses, as much is likely to change.

**Lived Experience Commentary: “All In This Together?”**

**By Beverley Chipp and Jo Lomani with contributions from Sarah Carr, Prisha Shah, and Nick Barber**.

This study assimilated international and grey literature written in several languages. Despite the inclusion of a wide variety of sources, there is an absence of discrete minority group perspectives and sources focusing on the disproportionate impact of COVID-19 on BAME groups in particular. The synthesis touches on the denial of liberties of people with MH problems but research is yet to explore aspects of urgency and emotionality around this issue or the effects of this as a secondary response.

Deprivation of rights from a fear that people cannot adequately socially distance, reducing the number of clinicians required to admit people under the Mental Health Act and inequalities of treatment for those with mental health problems who have COVID is unacceptable and worthy of future scrutiny.

Safety relating to mental health environments was omitted. Given the challenges of segregation without the unethical use of sedation and solitary confinement, attention should be directed towards ward design to minimise contagion.

Regarding people’s ability to self-manage, it is unclear to what extent this can be framed as ‘resilience’ in circumstances with few other options, and what can be maintained without support. Others may only opt to self-manage from fear of infection or concern about being burdensome to an overwhelmed NHS.

Reported satisfaction with virtual consultations naturally omits the voice of those unable to participate, and so conclusions should be viewed with caution. Digital exclusion is real and complex.

Issues raised in the paper – a triple whammy of poorer service, loss of rights (both informal and state sanctioned e.g. Coronavirus Act) and the reduced access to advocacy or legal services also have an aggregate relationship. The complexity of this effect requires deeper qualitative research. Going forward, it is vital to understand the long-term mental health consequences that pandemics have on different intersections of society.

***This is an independently written perspective from lived experience contributed by some of the co-authors with relevant experience***.

## Data Availability

The data consists of documents in the public domain, analysed through the method of framework synthesis.

## Acknowledgements

This paper presents independent research commissioned and funded by the National Institute for Health Research (NIHR) Policy Research Programme, conducted by the NIHR Policy Research Unit (PRU) in Mental Health. The views expressed are those of the authors and not necessarily those of the NIHR, the Department of Health and Social Care or its arm’s length bodies, or other government departments.

## Declarations

### Conflicts of interest/Competing interests

SJ, AS, SO and SC are grant holders for the NIHR Mental Health Policy Research Unit. JEC holds grants from NHS England & NHS Improvement outside of the submitted work.

### Ethics approval

Not applicable – analysis of material in the public domain

### Authors’ contributions

SJ, AS, and SO contributed to the original study proposal. SC, PB, and LSR drafted the study protocol with revisions by SJ, AS, NM, BLT, SO, and some members of the COVID-19 Mental Health Policy Research Unit Group. PB, TS and LSR led the searching and screening processes, with contributions from members of the COVID-19 Mental Health Policy Research Unit Group. SJ, LSR, PB, and TS led on development of the two frameworks and JN created the online version. LSR led on the data indexing and charting with contributions from PB, JN, JR, HS, TS, FB, JEC, PS, JL, BC, AS, SC, NB, ZD, SO, BLT, and most members of the COVID-19 Mental Health Policy Research Unit Group. SJ, TS, LSR, PB, JN, BLT, FB, JEC, HS, JR, PS, SC mapped, interpreted data, and summarised the charted data. SJ and JN drafted the initial report. BC and JL wrote the Lived Experience Commentary with contributions from SC, PS, and NB. NM, SO, SC, AS, SJ, and JN provided subject expertise and methodological guidance. All authors contributed to consecutive drafts and approved the final report.

## Notes

### Competing Interest Statement

SJ (lead author) AS, SO and SC are grant holders for the National Institute for Health Research Mental Health Policy Research Unit. JEC holds grants from NHS England & NHS Improvement outside of the submitted work.

### Author Declarations

This is an analysis using framework syntheissof documents that are published and in the public domain: no ethical approval is needed.

